# Negative association between higher maternal body mass index and breastfeeding outcomes is not mediated by DNA methylation

**DOI:** 10.1101/2023.11.01.23297893

**Authors:** Hannah R. Elliott, Chloe L. Bennett, Doretta Caramaschi, Sinead English

## Abstract

The benefits of breastfeeding for the health and wellbeing of both infants and mothers are well documented, yet global breastfeeding rates are low. One factor associated with low breastfeeding is maternal body mass index (BMI), which is used as a measure of obesity. The negative relationship between maternal obesity and breastfeeding is likely caused by a variety of social, psychological, and physiological factors. Maternal obesity may also have a direct biological association with breastfeeding through changes in maternal DNA methylation. Here, we investigate this potential biological association using data from a UK-based cohort study, the Avon Longitudinal Study of Parents and Children (ALSPAC). We find that pre-pregnancy body mass index (BMI) is associated with lower initiation to breastfeed and shorter breastfeeding duration. We conduct epigenome-wide association studies (EWAS) of maternal BMI and breastfeeding outcomes and candidate-gene analysis of methylation sites associated with BMI identified via previous meta-EWAS. We find that DNA methylation at cg11453712, annotated to PHTP1, is associated with maternal BMI. From our results, neither this association nor those at candidate-gene sites are likely to mediate the link between maternal BMI and breastfeeding.

## Introduction

Breastfeeding is associated with a multitude of benefits to mothers and infants, both in the short and long term, including reduced risk of breast cancer, diabetes and obesity, and improved cognition^1,2^. The World Health Organisation recommends exclusive breastfeeding (EBF) as the main infant feeding method for the first 6 months of life^3^. Despite health benefits, rates of breastfeeding initiation and duration remain low globally. A 2010 study on breastfeeding rates in the UK showed that, while 83% of mothers initiate breastfeeding, only 24% were EBF at 6 weeks, and 1% at 6 months^4^. These trends are apparent in countries with both high and low income, with evidence suggesting that increasing breastfeeding rates worldwide could lead to prevention of 823,000 deaths of children under 5 years old annually^2^. Strategies are required to increase breastfeeding rates, but for these to be most effective, understanding the mechanisms underlying reduced breastfeeding rates is essential.

Of several factors associated with reduced breastfeeding, maternal obesity is of particular concern. The prevalence of overweight and obesity among women of reproductive age is increasing globally: in 2016, it was estimated that 40% of women aged 18 or over were overweight^5,6^. Body mass index (BMI) denotes the weight of an individual relative to the square of their height and is used to categorise overweight and obesity^5,6^. Extensive studies have shown that obesity or high pre-pregnancy BMI are associated with lower breastfeeding initiation and shorter breastfeeding duration^6–9^. Potential causes behind this relationship include socio-cultural factors such as low self-esteem and body confidence, physical factors such as difficulties for the infant when latching to the breast, and physiological factors such as hormone imbalances or low milk supply in women who are overweight or obese^10–12^. Indeed, there is evidence that the second stage of lactogenesis (i.e., from 3 days up to one week after birth) is delayed in overweight women^13,14^. While the link between BMI and psychosocial factors associated with breastfeeding has been well explored^12,15^, there has been less investigation into the underlying biological or physiological mechanisms.

Here, we explore the biological association between high maternal BMI and reduced breastfeeding, focussing on the potential role of DNA methylation. DNA methylation is typically measured as attachment of a methyl group to a cytosine adjacent to a guanine nucleotide (referred to as a CpG site). It is a biological mechanism involved in regulation of gene expression that is influenced by both genetics and environment^16,17^. There is growing evidence from several cohort studies that there are maternal epigenetic differences associated with increased BMI^18–28^, with loci in biological pathways involved in lipid metabolism, adipose tissue hypoxia, and inflammation, although the relevant loci are not always consistent across studies. For most sites identified, causal inference analyses indicate that the likely pattern is that increased BMI leads to changes in methylation rather than vice versa^21,23,26^. Breastfeeding has been associated with differential methylation profiles in offspring^29,30^. Studies have also addressed how the DNA methylation profile of mothers changes across pregnancy^31^, with differential methylation in loci associated with metabolism and mammary gland development. Less understood, however, is how maternal DNA methylation is associated with breastfeeding outcome, and how this might depend on maternal BMI. For example, the physiological effect of delayed lactogenesis among overweight mothers may have an epigenetic association.

Here, we analyse data from a UK population study, the Avon Longitudinal Study of Parents and Children (ALSPAC), to investigate, first, the extent to which maternal BMI is associated with breastfeeding outcomes, and second, whether any such associations are potentially mediated by BMI-associated methylation in pregnant mothers. To achieve the latter, we conducted epigenome-wide association studies (EWAS) of methylation and its association with maternal BMI, breastfeeding initiation, and breastfeeding duration.

We also conducted specific candidate-gene analysis using a predetermined set of CpG sites known to be associated with BMI in adults, identified using a large meta-analysis of 18 studies ^28^, to ascertain whether these sites are also associated with BMI in pregnant women and whether methylation at these sites is associated with breastfeeding practices. Any associations found between the traits of interest and DNA methylation within the maternal epigenome could be used to support the hypothesis that DNA methylation acts as a mediator in the relationship between increased maternal BMI and reduced breastfeeding rates.

## Results

### Sample characteristics

The descriptive statistics generated for the ALSPAC sample used in our study, and ARIES subset are shown in Table 1. ARIES mothers are broadly similar to the ALSPAC sample, in line with previous evidence^32^. Even though the comparisons are generally consistent, ARIES mothers have a lower prevalence of smoking during pregnancy; and a higher proportion of ARIES mothers initiated and maintained breastfeeding practice compared to mothers in the broader ALSPAC dataset. We only included mothers with non-missing information in the EWAS. As such, 724 samples were included in the EWAS of maternal BMI, 718 samples were included in the EWAS of breastfeeding initiation and 602 individuals were included in the EWAS of breastfeeding duration.

**Table 1:** Comparison of the baseline characteristics for the exposure variables and covariates used in the EWAS between mothers included in ALSPAC only and the ARIES subset.

| **Variable** | **ALSPAC dataset**<br>*Mean (standard deviation) OR %* | **ARIES subset (n=933)**<br>*Mean (standard deviation) OR %* |
| --- | --- | --- |
| Pre-pregnancy BMI (continuous) | **n = 11376**<br>22.9 (3.9) | **n = 861**<br>22.8 (3.6) |
| Pre-pregnancy BMI (categorical) | **n = 11376** | **n = 861** |
| <25 | 79.33% | 82.69% |
| 25-29 | 15.10% | 12.78% |
| >= 30 | 5.56% | 4.53% |
| Parity | **n = 12774** | **n = 902** |
| Nulliparous | 44.97% | 46.90% |
| Multiparous | 55.03% | 53.10% |
| Age at delivery (years) | **n = 11539**<br>28.3 (4.8) | **n = 898**<br>29.6 (4.3) |
| Smoked during pregnancy | **n = 12912** | **n = 916** |
| Yes | 18.79% | 9.28% |
| No | 81.21% | 90.72% |
| Education level | **n = 11413** | **n = 894** |
| Below A-Level | 63.36% | 48.55% |
| A Level and above | 37.64% | 51.45% |
| Occupation | **n = 9855** | **n = 820** |
| Manual | 19.90% | 13.29% |
| Non-manual | 80.10% | 86.71% |
| Breastfeeding intention | **n = 11694** | **n = 890** |
| No | 19.68% | 10.22% |
| Maybe | 14.19% | 12.81% |
| Yes | 66.13% | 76.97% |
| Breastfeeding initiation (ever breastfed) | **n = 12035** | **n = 912** |
| Yes | 77.25% | 89.58% |
| No | 22.75% | 10.42% |
| Breastfeeding duration (categorised) | **n = 10875** | **n = 887** |
| Never | 24.51% | 13.19% |
| Up to 3 months | 31.99% | 28.18% |
| Up to 5 months | 12.83% | 16.35% |
| 6 or more months | 30.67% | 42.28% |
| Breastfeeding duration (months) | **n = 7862**<br>5.7 (4.5) | **n = 754**<br>6.4 (4.4) |

### Association between maternal BMI and breastfeeding (initiation and duration)

In both univariate and multivariate models (including potentially confounding variables of breastfeeding intention, maternal smoking, age, occupation, parity, and education), maternal BMI was negatively associated with whether a mother initiated breastfeeding or not (univariate model, n=10,548; odds ratio OR [95% confidence interval CI]: 0.962 [0.951, 0.973]; multivariate model, n= 7,704; OR [CI]: 0.944 [0.921, 0.969]). Similarly, among mothers who did breastfeed, the duration of breastfeeding was lower for those mothers with higher BMI (univariate model, n= 7,166; Beta [CI]: −0.108 [−0.138, −0.079]; multivariate model, n= 5,645; Beta [CI]: −0.091 [−0.123, −0.059]). When we analysed breastfeeding duration as a categorical variable, we also find that higher BMI is associated with shorter breastfeeding outcomes (univariate model, n= 9,761; OR [CI]: 0.946 [0.936, 0.954; multivariate model, n= 7,298; OR [CI]: 0.944 [0.931, 0.956]). For results describing all covariates, see Tables S1–3 (ESM). Note that we found qualitatively similar results when we considered categorised pre-pregnancy BMI (rather than continuous) as a covariate, and when we analysed breastfeeding duration (in months) as a time-to-event process, with proportional hazards models (Tables S4, Figure S1, ESM).

### Epigenome-wide association analyses

#### (i) Maternal BMI

In the univariate model, three CpG sites were identified below the genome wide threshold (*p*< 2.4 x 10^−7^) when maternal BMI was modelled as the exposure variable. A total of 45 CpG sites were associated with maternal BMI at *p*< 1.0 x 10^−5^. A lambda of 2.34 indicated moderate inflation of *p-*values. In multivariate analysis, one CpG site was identified below the genome wide threshold (*p*< 2.4 x 10^−7^) when maternal BMI was modelled as the exposure variable (see Figure 1). The lambda value (λ = 1.29) indicates minor inflation of the results. A total of 20 CpG sites were associated with maternal BMI at *p*< 1.0 x 10^−5^ in the multivariate analysis, as detailed in Table 2.

**Figure 1.**
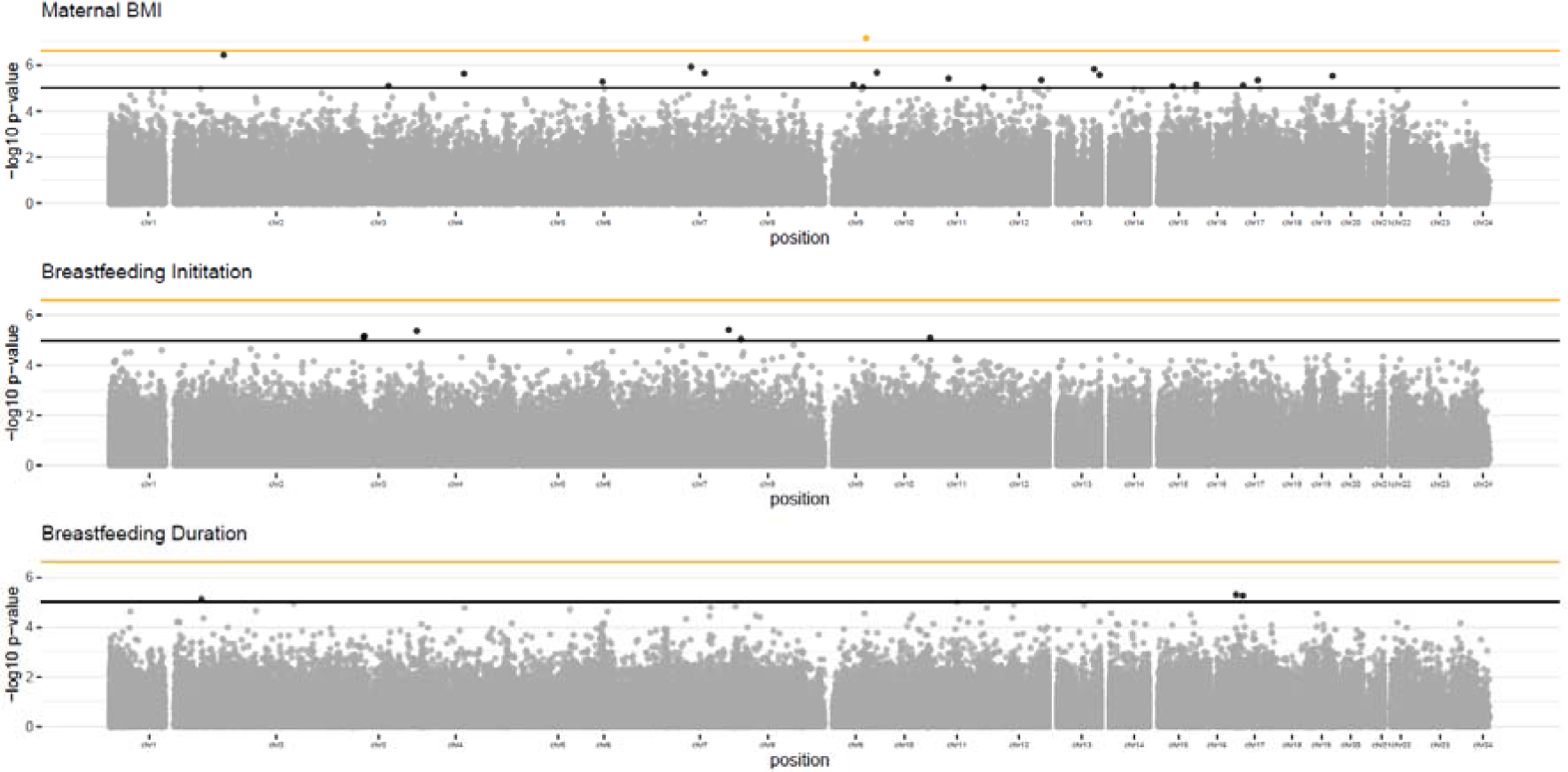
Manhattan plots showing the association between DNA methylation and phenotype (maternal BMI, breastfeeding initiation, and breastfeeding duration) measured in maternal blood samples from the ALSPAC cohort. Each point on the plots represents an individual CpG site. The orange threshold line represents a genome wide threshold of p< 2.4 x 10^−7^ and CpG sites with p-values below this threshold are highlighted in orange. The black threshold line represents a relaxed threshold of p<1.0 x 10^−5^ and CpG sites with p-values below this threshold are highlighted in black.

**Table 2.**
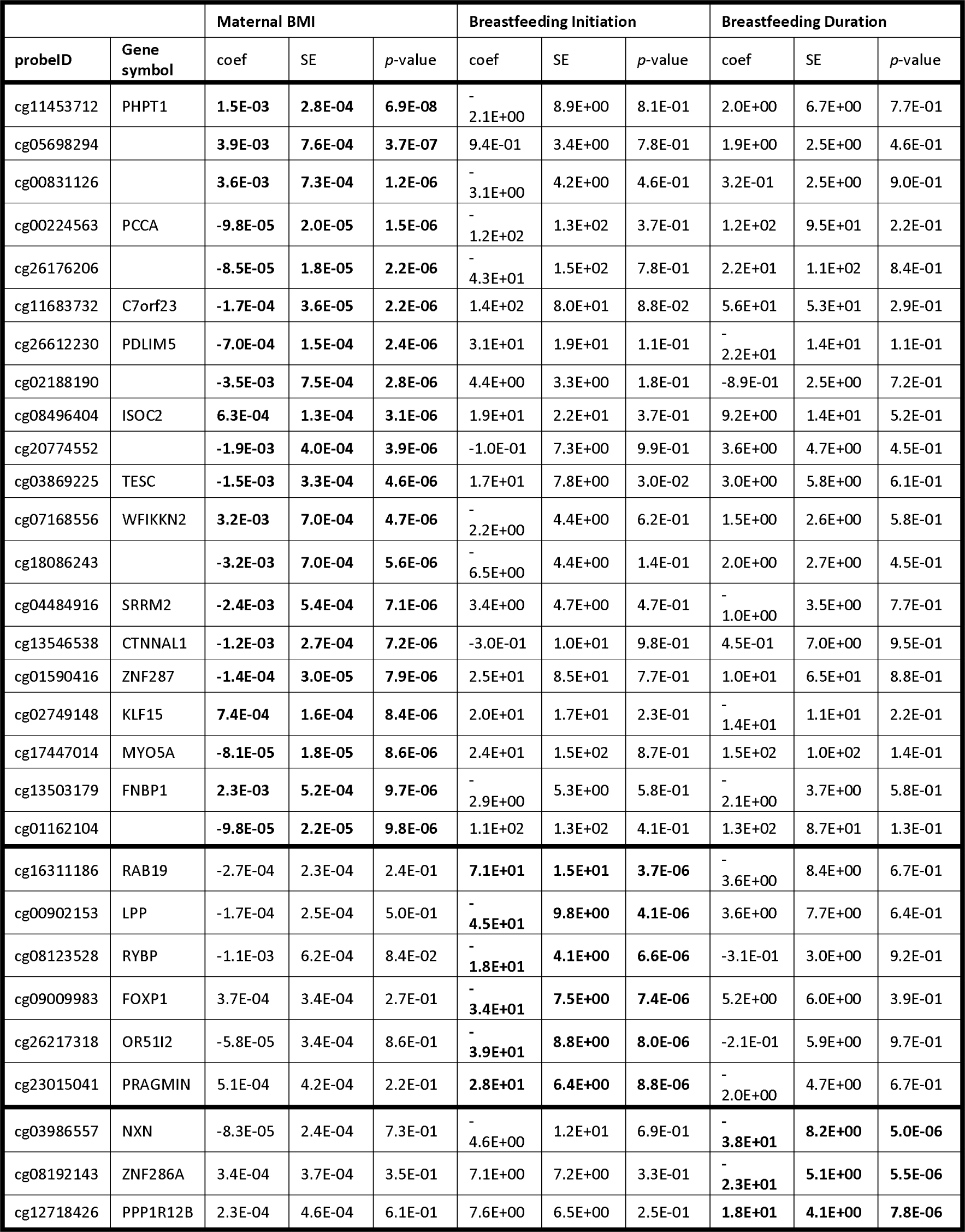
Table showing the associations between DNA methylation and maternal phenotype for fully adjusted EWAS at p<1.0 x 10^−5^ across the three EWAS conducted in ALSPAC.

#### (ii) Breastfeeding initiation

In the univariate model, we did not identify any CpGs associated with breastfeeding initiation at the genome wide threshold (*p*< 2.4 x 10^−7^) and only one CpG at *p*< 1.0 x 10^−5^. A lambda value of 1.33 indicated minor inflation of *p-*values. In the multivariate model, we did not identify any CpG sites associated with breastfeeding initiation below the genome wide threshold (*p*< 2.4 x 10^−7^, Figure 1). The lambda value (λ=1.27) indicates minor inflation of the results. Breastfeeding initiation was associated with 6 CpGs in the multivariate model at *p*< 1.0 x 10^−5^ (Table 2).

#### (iii) Breastfeeding duration

In the univariate model, we identified one CpG site associated with breastfeeding duration, coded as a continuous variable, below the genome wide threshold (*p*< 2.4 x 10^−7^). Three CpG sites were at *p*< 1.0 x 10^−5^. The lambda value (λ=1.00) indicated no inflation of *p-* values. In multivariate analysis, we did not identify any CpG sites associated with breastfeeding duration below the genome wide threshold (*p*< 2.4 x 10^−7^, Figure 1). The lambda value (λ=1.02) also showed no genome wide inflation of *p-*values versus those expected. Continuously measured breastfeeding duration was associated with 3 CpG sites in the multivariate model at *p*< 1.0 x 10^−5^ (Table 2).

Full EWAS results for all EWAS presented in this paper are detailed in Supplementary Tables S5-7, ESM and Manhattan plots of fully adjusted models are shown in Figure 1.

### Candidate gene analysis

The results for the candidate gene analysis found that none of the 52 CpG sites drawn from the Do *et al*. meta-EWAS analysis of BMI ^28^ met the *p-*value threshold of 1.00 x 10^−05^ (Table 2) in ALSPAC EWAS of BMI, breastfeeding initiation or breastfeeding duration. ALSPAC test statistics for the CpG sites identified in Do *et al.* are shown in Supplementary Table 8, ESM. We evaluated the consistency in direction of effects between our EWAS of maternal BMI and results reported by Do *et al.* and calculated the proportion of CpGs which had the same direction of effect using a binomial test of the null hypothesis that the proportion is equal to 0.5. 39/52 CpG sites showed consistency of direction of effect estimate, binomial test *p-* value 4.1 x 10^−4^. We therefore demonstrate that there is weak evidence for association between BMI and DNA methylation in pregnant women in the ALSPAC cohort, but effect estimates are consistently in the same direction as previously reported in the literature. None of the CpG sites identified to be associated with BMI in the Do *et al.* study are, however, associated with the breastfeeding measures assessed in ALSPAC.

## Discussion

In this study, we first confirmed previously established associations between high maternal pre-pregnancy BMI and lower initiation and duration of breastfeeding, using analysis of the UK ALSPAC population study. We then conducted EWAS to identify whether maternal BMI and breastfeeding practice were associated with DNA methylation at CpG sites in the maternal genome, which could act as potential mediators in the relationship between increased maternal BMI and lowered breastfeeding rates. We found BMI to be associated with DNA methylation at one CpG site and putatively associated with further 19 CpG sites. However, none of these sites were associated with breastfeeding initiation or duration.

We find a negative association between higher maternal BMI and breastfeeding outcome – both in terms of whether it was initiated and how long it lasted – in line with several other studies on this topic across diverse cohorts^6,8,11^. Given that obesity rates are rising rapidly across the world, and breastfeeding rates remain low despite known benefits, studies investigating why high-BMI mothers are less likely to breastfeed their babies could help inform policy to improve breastfeeding outcome. We acknowledge limitations of using BMI as a measure of obesity^33^, yet appreciate that it is a practical measure and can be used at scale in population studies such as ALSPAC.

Potential factors explaining the replicated negative relationship between BMI and breastfeeding outcome include delayed lactogenesis. The second stage of lactogenesis, which occurs on average around 3 days up to a week after birth, is more likely to be delayed in overweight women^13,14^. We originally hypothesised that biological factors behind this phenomenon could be captured by epigenetic biomarkers in peripheral blood during pregnancy. Finding such biomarkers could have implications in identifying women in need of extra support and could further our understanding of the mechanisms behind delayed lactogenesis. However, we did not find strong evidence to support our hypothesis. Other potential factors that have been previously linked with suboptimal breastfeeding outcomes are parity, delivery method, experience of epidural, and infant behaviour after birth^13,14^. We adjusted our analyses for parity as a potential confounder in the associations between maternal BMI and breastfeeding. Further studies should explore the other factors as potential mediators between high BMI and low breastfeeding initiation and duration.

A robust relationship between BMI and DNA methylation was evidenced by previous large epigenome-wide association studies in the general population^23,28^, therefore we hypothesised that we would find a similar link in UK pregnant women. Although less powered, our study found associations in the same directions as found by the largest EWAS of BMI^28^. We also found novel associations, though we are cautious about these unless replicated in other cohorts. The strongest association is located on the PHTP1 gene, encoding a phosphatase not previously linked to obesity. To our knowledge, this is the first study investigating this question in pregnant women, and our results suggest that during pregnancy the relationship between BMI and DNA methylation might be altered, potentially due to immune-metabolic changes typical of pregnancy^34,35^. Previous studies that attempted to investigate the link between BMI and DNA methylation during pregnancy focussed on candidate genes encoding leptin and adiponectin and found associations^36^. Neither of these loci appear on our top list of genes associated with BMI, however, using the relaxed *p-*value threshold.

Our EWAS of breastfeeding outcome did not identify CpG sites whose methylation in the peripheral blood of pregnant women is associated with breastfeeding outcomes in our study. To our knowledge, this is the first study examining this link and we conducted an epigenome-wide scan across more than 450,000 CpG sites. It is possible that if there are true associations, their effect sizes will be too small to detect with our sample. The lack of study populations with DNA methylation data on pregnant women limited our sample size. However, if the top associations found with breastfeeding using the relaxed threshold are true, they suggest implications of the LPP and NXN genes. Methylation at these genes in the offspring at birth was also associated with maternal BMI and pre-pregnancy overweight/obesity in a previous study, though at different CpG sites in the same loci^37^. DNA methylation at these genes was not affected antenatally in the mothers in our study. One explanation for this is an intergenerational effect via alterations of fetal development and with consequent challenges around delivery and in the baby’s feeding behaviour, rather than the mother’s own ability to lactate. For instance, a recent study shows that early pregnancy BMI is linked with DNA methylation in placental tissue^38^.

We also conducted a targeted approach, analysing candidate CpG sites previously found to be associated with BMI from a broader EWAS ^28^. Similar to our EWAS analysis, we did not find support that methylation associated with BMI is linked with reduced breastfeeding outcomes. These results, together with the EWAS results, do not support our hypothesis that DNA methylation is implicated in the link between maternal BMI and breastfeeding practice. It is possible, however, that biological mechanisms unrelated to peripheral blood DNA methylation are involved in mediating the link between BMI and reduced breastfeeding. Further studies should be focused on elucidating these mechanisms. Another explanation is that biological mechanisms are less involved and that social and cultural aspects play a stronger role in the negative association between maternal overweight and breastfeeding outcome. For instance, many women fail to reach their own breastfeeding goals because of discouraging factors such as employer support, husband involvement and social attitudes^39^. Moreover, studies have shown there can be a general negative public attitude towards breastfeeding^40^, and it is possible that overweight women are more vulnerable to these attitudes^41^. In any case, considering the challenges that overweight women face around breastfeeding, more dedicated support might be beneficial to increase breastfeeding rates.

Despite the main strengths of our study accessing a large and rich intergenerational dataset and exploring a novel question, our study has several limitations. First, we had a small sample size for the methylation studies. Although the initial ALSPAC cohort recruited more than 14,000 pregnancies, the subsample with antenatal blood samples and data on BMI, breastfeeding outcome and covariates data is lower than 1000 mother-child pairs. This is considered an under-powered sample size for EWAS studies of outcomes such as BMI and it is increasingly common to combine multiple cohorts to greatly increase the number of subjects. For example, Sharp et al^37^ were able to meta-analyse the association between maternal pre-pregnancy BMI and methylation in the peripheral blood of offspring across 19 longitudinal cohorts, resulting in a sample size of 9,340. Obtaining a sample size of this magnitude increases internal and external validity and reliability of results, as natural variation can greatly affect DNA methylation and hence the outcomes of the study^42^. A second drawback to our study was that the analyses were not replicated. Future similar studies should be conducted to further investigate the association between increased BMI and DNA methylation in antenatal blood and how this methylation is associated with breastfeeding practice. An additional limitation of the EWAS in this study is tissue availability. The advantage of using peripheral blood as a surrogate tissue is that it can be collected in large quantities and is easy to store. In the context of this study, to assess the association between breastfeeding practice and changes to the DNA methylation profile, breast tissue samples (preferably prenatally or during the period where the mother breastfeeds) would be the optimal sample tissue. This would lead to more accurate and compelling analyses of associations between breastfeeding practice and breast tissue DNA methylation.

In conclusion, this study provides the first insight into the maternal epigenome and its association with maternal pre-pregnancy BMI and breastfeeding initiation and duration. The results from this study did not support our mediation hypothesis, that DNA methylation could explain putative pattern of reduced breastfeeding among overweight mothers, yet owing to our underpowered study we are cautious to infer that this does not mean that BMI-associated methylation is not associated with reduced breastfeeding at all. Our novel investigation into the relationship between maternal exposures such as BMI and breastfeeding on maternal DNA methylation provides the framework for more in-depth study into the physiological mechanisms that impede a mother’s ability to breastfeed, and future studies could expand our analyses to more cohorts, and consider other biological pathways.

## Methods

### Study cohort

The Avon Longitudinal Study of Parents and Children (ALSPAC) is a birth cohort study that investigates modifiable influences on child health and development^43,44^. Pregnant women resident in Avon, UK, with expected dates of delivery between 1st April 1991 and 31st December 1992 were invited to take part in the study. The initial number of pregnancies enrolled was 14,541. Of the initial pregnancies, there was a total of 14,676 foetuses, resulting in 14,062 live births and 13,988 children who were alive at 1 year of age.

### Ethical approval

Ethical approval for the study was obtained from the ALSPAC Ethics and Law Committee and the Local Research Ethics Committees. Informed consent for the use of data collected via questionnaires and clinics was obtained from participants following the recommendations of the ALSPAC Ethics and Law Committee at the time. Consent for biological samples has been collected in accordance with the Human Tissue Act (2004).

### Study measures

#### (i) Maternal body mass index (BMI)

BMI is the metric currently used for defining anthropometric height/weight characteristics and typically categorizing them into groups, and its common interpretation is that it is a measure of extent of overweight and obesity^5,6^. We used self-reported measures of pre-pregnancy weight (in kilograms) and height (in metres) from the ‘About Yourself’ mother-completed questionnaire which can be found in the ALSPAC data dictionary. Maternal BMI was calculated as weight (kg) / height (m^2^) as a continuous variable; and analyses were repeated with maternal BMI categorised as D30 (0) or ≥30 (1), following standard WHO categorisation^45^ which defines obese as ≥30, overweight as 25-29 and ‘normal’ or ‘acceptable’ as <25.

#### (ii) Breastfeeding outcome

Data regarding breastfeeding outcome were collected from two questionnaires administered to the mother when the child was 6 months and 15 months of age respectively. Three different measures of breastfeeding were used: (1) breastfeeding initiation, a binary indicator (‘yes’ or ‘no’) of whether the mother had ever initiated any breastfeeding before the child had reached 6 months of age; and, for those mothers who initiated breastfeeding, (2) continuous breastfeeding duration, measured as the age in months (from 0–15) of the child when breastfeeding stopped, derived from the 15-month questionnaire, and (3) categorical breastfeeding duration, i.e. never, up to 3 months, 3 to 5 months and 6 or more months, derived from the 6-month questionnaire, as this provided data for a larger number of mothers. Further details on how these variables were derived can be found in the ‘My Son/My Daughter’ mother-completed questionnaire in the ALSPAC data dictionary.

#### (iii) DNA methylation

DNA methylation data was obtained using Illumina 450k BeadChip arrays from the Accessible Resource for Integrated Epigenomics Studies (ARIES) project^32^. Blood samples from a sub-set of 1,018 ALSPAC participants were taken at three time points for the child (neonatal, childhood (mean age 7.5 years) and adolescence (mean age 17.1 years)) and at two time points for the mothers (antenatal and during their child’s adolescence (mean age 17.1 years)). The DNA methylation data used in this study were from samples collected at the antenatal time point from mothers.

Normalisation was carried out using the *meffil* R package^46^. Samples showing evidence of population stratification based on ALSPAC genetic data were removed (n=29). Methylation status was quantified as the ratio of the methylated probe intensity and the overall intensity (sum of methylated and unmethylated probe intensities) resulting in a beta-value between 0 (completely unmethylated) and 1 (completely methylated)^47^. This beta-value represents the proportion of methylated cells within the sample. The impact of outliers was reduced by setting all methylation data points outside 3 times the interquartile range from the 25^th^ to the 75^th^ percentiles as missing. Final EWAS analyses included a total of *N =* 482,855 probes and 933 individuals.

#### (iv) Covariates

The following variables were included in the analyses to adjust for confounding effects^48–51^. Maternal age at delivery was derived from a questionnaire administered 8 weeks after the child’s birth using the self-reported date of birth of the mother and the date of birth of the child, measured as a continuous variable. Parity was categorised as either nulliparous or multiparous derived from a questionnaire administered at 18 weeks’ gestation. Maternal smoking during pregnancy was categorised as either never smoking during pregnancy or smoking during pregnancy, derived from a questionnaire administered at 23 weeks’ gestation. The following variables were derived from a questionnaire administered at 32 weeks’ gestation: whether the mother intended to breastfeed in the first month (recategorized as ‘yes’, ‘maybe’ or ‘no’ following methodology of Jones et al.^52^); maternal education, categorised as having completed qualifications below A-Levels or completed the qualifications of A-Levels and above; and occupational social class, categorised according to whether the mother or father had a manual or non-manual occupation (whichever was highest) according to the National Statistics Socio-economic Classification. For the methylation analyses, estimated cell counts were derived^53^ and included as model covariates. Given the low ethnic diversity in our sample (<3% identified as non-white), we did not include maternal ethnicity as a covariate.

### Statistical analysis

Analyses were carried out in R (version 4.1.0 ^54^). Descriptive statistics were calculated for the ALSPAC sample and for the ARIES subset. Any mothers with missing data for any of the exposure or covariates were removed, leaving only complete cases for the primary analyses. The final samples contained only singleton births. Descriptive statistics for each subset (both for the ALSPAC phenotype dataset and ARIES subset EWAS sample) were subsequently calculated, to check for similarity of samples across analyses.

#### (i) Association of maternal BMI with breastfeeding

We investigated whether maternal BMI was associated with breastfeeding outcome (i.e., initiation and duration) using multiple regression. For each analysis, we first ran a univariate model considering only the association between maternal BMI and the breastfeeding measure of interest. We subsequently repeated the analysis including covariates known previously to be associated with breastfeeding (intention to breastfeed, maternal smoking, age, occupational social class, parity, and education). For analysis of breastfeeding initiation, we used a logistic regression with binomial error structure; for analysis of continuous breastfeeding duration, we used a linear model with gaussian error; and for analysis of categorical breastfeeding duration, we used ordinal logistic regression. We also conducted analyses on continuous breastfeeding duration as a time-to-event analysis using proportional hazards models, using the function survfit for univariate analyses and coxph for multivariate, from the ‘survival’ package in R^55^.

#### (ii) Epigenome-wide association study (EWAS)

Three EWASs were carried out using multiple linear regression to estimate the association of DNA methylation in maternal antenatal peripheral blood with maternal BMI (*N =* 724), breastfeeding initiation (*N =* 718), and breastfeeding duration as a continuous measure *(N* = 602). For the EWAS of maternal BMI, we modelled DNA methylation as the outcome. For EWASs of breastfeeding, we modelled breastfeeding as the outcome. For analysis of breastfeeding initiation, we used a logistic regression with binomial error structure; for analyses of continuous breastfeeding duration and maternal BMI, we used a linear model with gaussian error. All models were adjusted for maternal age, parity, maternal smoking behaviour, maternal education, occupational social class, intention to breastfeed, cell-type estimates and 10 Surrogate Variables generated using Surrogate Variables Analysis to remove unmeasured confounding^56^. For the EWASs on breastfeeding, maternal BMI was also included as a covariate, to account for potential direct effects of BMI on breastfeeding (in addition to those mediated through methylation).

To control for multiple testing, we set the genome-wide threshold at *p*< 2.4 x 10^−7^ ^57^ as well as reporting results at a relaxed threshold of *p*<1.0 x 10^−5^. For each EWAS, the genomic inflation factor (lambda λ) was calculated to quantify the extent of the inflation and the excess false positive rate; quantile-quantile (Q-Q) plots were used as a visual tool to mark deviations of the observed distributions from the expected distribution of *p-*values. EWASs were carried out using R package *ewaff* (version: 0.0.2; https://github.com/perishky/ewaff).

#### (iii) Candidate-CpG analysis

To further investigate the potential association between maternal BMI and DNA methylation and whether methylation is subsequently associated with breastfeeding practice, a candidate gene analysis was carried out using a recent meta-EWAS of adult BMI across 18 studies by Do *et al.*^28^. In this analysis, we restricted ALSPAC EWAS results to consider only CpG sites that were associated with BMI in the Do *et al.* study, which identified 52 CpG sites passing a false discovery rate adjustment in their meta-EWAS of adult BMI. We then investigated whether these specific CpG sites were associated with maternal BMI and breastfeeding practice in ALSPAC. We used a binomial test to assess agreement in direction of effect estimates between the BMI meta-EWAS of Do *et al.* and the EWAS of BMI in ALSPAC.

## Supporting information

ESM_Figure

ESM_Tables

## Acknowledgements

We are extremely grateful to all the families who took part in this study, the midwives for their help in recruiting them, and the whole ALSPAC team, which includes interviewers, computer and laboratory technicians, clerical workers, research scientists, volunteers, managers, receptionists and nurses.

## Author contribution

C.B. designed the study with input from D.C., S.E. and H.R.E.; C.B. conducted the initial analyses, S.E. (phenotype analysis), H.R.E. (EWAS) and D.C. (both) verified these and conducted the final analyses included in the report. All authors drafted the manuscript and provided comments and edits.

## Data Availability

The ALSPAC study website contains details of all the data that is available through a fully searchable data dictionary and variable search tool http://www.bristol.ac.uk/alspac/researchers/our-data/. ALSPAC data is available on request by application to the ALSPAC executive committee. The ALSPAC data management plan (available here: www.bristol.ac.uk/alspac/researchers/access/) describes in detail the policy regarding data sharing, which is through a system of managed open access.

