## Supplementary material for "Negative association between higher maternal body mass index and breastfeeding outcomes is not mediated by DNA methylation": ESM_Figure

Supplementary information for: “**Negative association between higher maternal body mass index and breastfeeding outcomes is not mediated by changes in DNA methylation”**

**Figure S1.** Plot from univariate survival analysis of cessation of breastfeeding for mothers of different BMI categories.

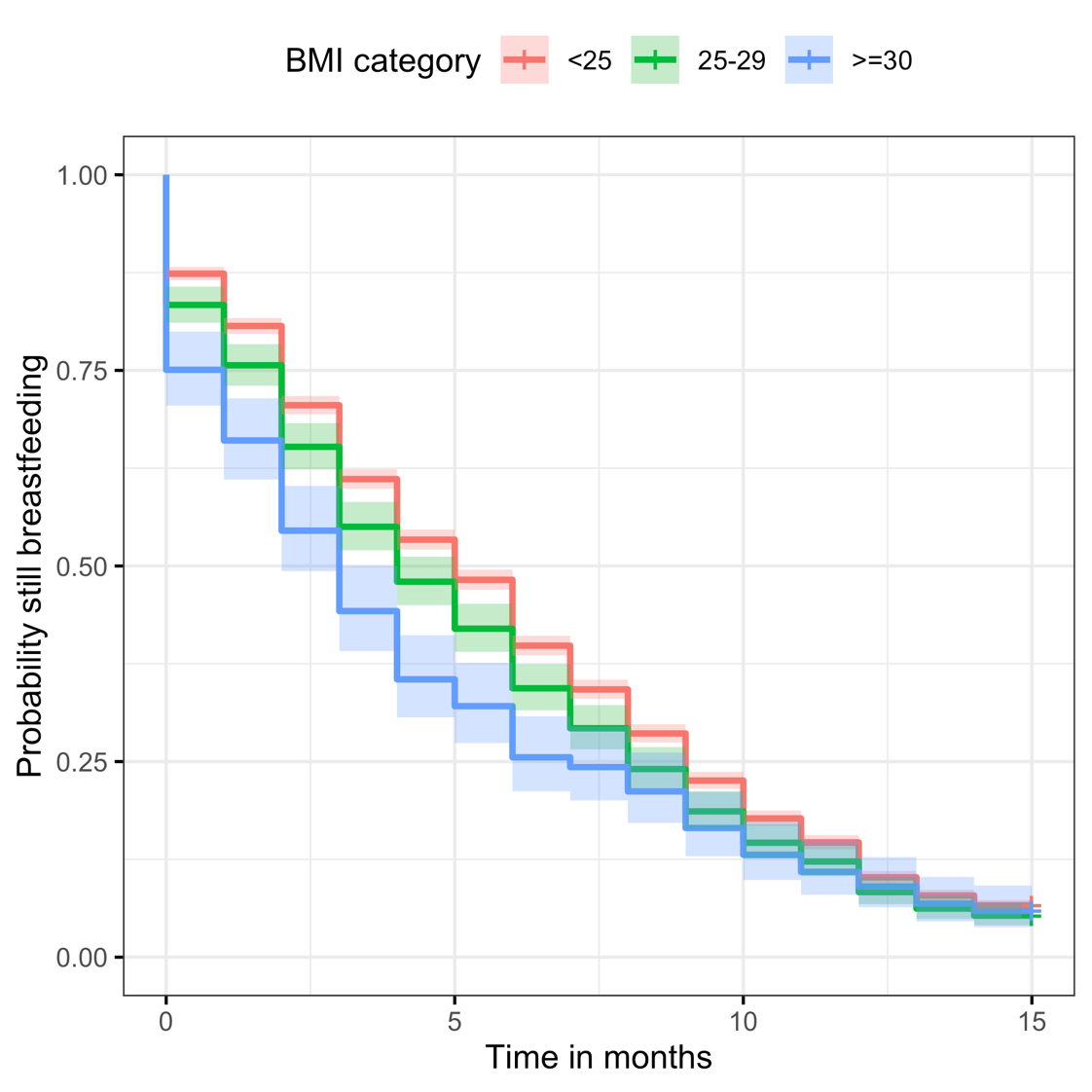
